# Life’s Essential 8, Genetic Susceptibility, and Incident Ischemic Stroke: A Prospective Cohort Study

**DOI:** 10.1101/2025.02.28.25323130

**Authors:** Runsheng Yang, Zhongmin Yin, Jia Luo, Jingjing Wang, Weijing Wang, Dongfeng Zhang

## Abstract

**BACKGROUND:** Life’s Essential 8 (LE8) is an important indicator to assess cardiovascular health (CVH), yet its association with ischemic stroke (IS) remains unclear. We aimed to investigate the association of LE8 with IS, as well as the underlying inflammatory mechanisms. Furthermore, we also aimed to explore whether genetic susceptibility moderated this association.

**METHODS:** A total of 202,215 participants aged 40-69 from the UK Biobank were analyzed between 2006 and 2010, with follow-up conducted until 2022. The American Heart Association’s LE8 framework was used to calculate the LE8 scores. The link between LE8 scores and IS risk was assessed using Cox proportional hazards models. 1000 non-parametric bootstrapping simulations were used to calculate the mediation effect. CVH levels and genetic susceptibility were classified as high, medium, and low based on their LE8 scores and polygenetic risk scores (PRS), respectively.

**RESULTS:** 2,515 individuals experienced IS over a median follow-up period of 13.73 years. In comparison to the low CVH group, the medium CVH group exhibited a 26% reduction in the risk of IS (HR 0.74; 95% CI, 0.65-0.80), while the high CVH group demonstrated a 53% reduction (HR 0.47; 95% CI, 0.39-0.57). An increased LE8 scores were inversely correlated with the risk of IS (HR: 0.84; 95% CI: 0.80-0.87), partially mediated by inflammatory markers. According to the joint effect analysis, individuals with low genetic susceptibility and high CVH had the lowest IS risk (HR: 0.20; 95% CI: 0.13-0.30) when compared to those with genetic susceptibility and low CVH.

**CONCLUSIONS:** The findings imply that higher LE8 scores and CVH levels are associated with a lower incidence of IS, partially mediated by inflammatory markers. This protective relationship can be strengthened in cases of lower genetic susceptibility.

## INTRODUTION

Stroke is a predominant cause of death and disability worldwide^1^. The most common form of stroke is ischemic stroke (IS), which accounts for approximately 71% of all strokes^1,2^. The IS refers to the infarction of the brain, spinal cord, or retina due to the occlusion of cerebral arteries^3^. In 2021, IS affected around 70 million individuals, establishing itself as a significant global health challenge^4^. Therefore, recognizing the risk factors for IS is crucial to managing and preventing it.

Cardiovascular health (CVH) was defined by the American Heart Association (AHA) in 2010 and assessed using the Life’s Simple 7 (LS7) score system^5^. Research have shown that LS7-measured CVH is correlates negatively with several cardiovascular and non-cardiovascular illnesses^6–8^. In 2022, the AHA updated the CVH evaluation system in response to the shortcomings of the LS7 classification, which resulted in the creation of Life’s Essential 8 (LE8)^9^. The revised framework encompasses eight metrics that assess both behavior (sleep health, nicotine exposure, physical activity and diet) and biological factors (non-high-density lipoprotein (non-HDL) cholesterol, blood pressure, blood glucose, and body mass index (BMI))^9^. LE8 incorporates a sleep assessment and refines the scoring criteria for indicators such as smoking and cholesterol. Furthermore, it optimizes the scoring system, with each indicator now ranging from 0 to 100, replacing LS7’s 0-2. Those refinements enhance LE8’s ability to capture subtle health changes, thereby enhancing its overall sensitivity and comprehensiveness^9^.

Although extensive research have investigated the relationship between LS7-measured CVH and incident IS^10–12^, studies examining CVH assessed using LE8 remain scarce. The Kailuan cohort in China reported a negative correlation between CVH assessed with LE8 and IS risk^13^. Another study indicated that no association exists between LE8 and IS in American Indian populations^14^. However, the findings of these two studies may be limited by the relatively short follow-up duration and small sample sizes. Furthermore, the mediating effects between LE8 scores and IS remains unclear. Studies have demonstrated an inverse relationship between Life’s Essential 8 metrics scores and C-reactive protein (CRP) levels^15^. Increased inflammation levels may result in the development of cardiovascular disorders, including IS^16,17^. Therefore, we hypothesize that higher LE8 scores may decrease the incidence of IS by lowering inflammatory markers. Moreover, genetic factors plays a crucial part in the occurrence of IS^18,19^. Studies indicate that lifestyle factors can interact with genetic susceptibility^20,21^. The likelihood of IS caused by genetic susceptibility to IS may be reduced by adopting a healthy lifestyle^21^. However, it remains uncertain whether the CVH evaluated by LE8 interacts with genetic susceptibility.

Therefore, we aimed to use data from the UK Biobank to investigate the relationship between CVH levels and subscale scores, as measured by LE8, with the risk of IS, as well as the underlying inflammatory mechanisms. Furthermore, whether this association could be influenced by genetic predisposition was also investigated.

## METHODS

### Study population

This study employed information from a prospective cohort study named the UK Biobank. Between 2006 and 2010, the study recruited approximately 500,000 individuals between the ages of 40 and 69 from 22 review centers across the United Kingdom^22^. The UK Biobank collected extensive genetic and non-genetic data regarding various diseases via interviews, questionnaires, physical activity assessments, and biological sample collection. Others have provided comprehensive descriptions of the research design and methodologies of the UK Biobank^22^. The North-West Multi-Centre Research Ethics Committee approved the UK Biobank study, and written informed consent was provided by all subjects.

This research excluded participants for the following reasons: absence of any components of LE8 (n=231,968), pregnancy at the study’s onset (n=73), loss to follow-up during the research duration (n=690), pre-existing cardiovascular disease at the study’s start (n=40,204), or missing covariate data (n=27,220). The primary analysis consisted of a total of 202,215 participants. Participants lacking mediator variables had been excluded (n=5,117), producing a final sample of 197,095 participants for analysis of mediation. Figure S1 presents the participant inclusion and exclusion flow chart. The STROBE (Strengthening the Reporting of Observational Studies in Epidemiology) reporting requirements were followed in this study (Supplementary STROBE checklist).

### Assessment of LE8 scores and CVH

The AHA guidelines require that the LE8 scores are derived from eight metrics, which encompass both behavior indicators (sleep health, nicotine exposure, physical activity and diet) and biological indicators (non-HDL cholesterol, blood glucose, blood pressure, and BMI)^23^. The LE8 scores, which also ranges from 0 to 100, is determined by taking the free of weight average of eight indications, each of which is assessed on a scale of 0 to 100. Each metric and the LE8 scores indicate improved health with an increase in the score. The dietary and nicotine exposure scores from the American Heart Association were modified to align with UK Biobank data^24^. All relevant metrics were assessed utilizing baseline data. According to the standards provided by AHA, the LE8 scores are classified into three separate categories: low CVH (ranging from 0 to 49), moderate CVH (ranging from 50 to 79), and high CVH (ranging from 80 to 100)^9^. Table S1-2 presents detailed assessment methods.

### Assessment of Ischemic Stroke

For the purpose of identifying IS, this study utilized the tenth International Classification of Diseases (ICD-10). The majority of the data was sourced from hospital inpatient records, with additional information obtained from death records and self-reports. The IS events were identified as diagnoses with ICD-10 code I63 that occurred from baseline to the ending of follow-up (October 31, 2022). The follow-up period began at enrollment in the UK Biobank cohort and continued until the censoring date, an IS, or death, whichever occurred first. Table S3 provides a detailed list of specific ICD-10 codes and their corresponding UK Biobank disease source codes.

### Assessment of Inflammatory Mediators

We collected data on inflammatory markers from blood tests performed on participants in the UK Biobank (visit https://biobank.ndph.ox.ac.uk/showcase/ukb/docs/haematology.pdf). This study performed an exploratory analysis to examine the influence of inflammation on the relationship between LE8 scores and IS, focusing on the mediating role of inflammatory indicators, including CRP, platelets, monocytes, white blood cell (WBC) count, and eosinophils. In the analysis, these markers underwent log transformation to approximate a normal distribution.

Alongside individual inflammation markers, we implemented a comprehensive assessment standard for chronic low-grade inflammation, referred to the INFLA-score. The combination of CRP, WBC count, platelet count, and the granulocyte/lymphocyte ratio (GrL) provides an extensive evaluation of an inflammatory status of individual^25^. To calculate the INFLA score, one must derive individual indicator scores from the quartiles (1 to 10) of the four indicators within the sample. Percentiles 1 to 4 receive negative scores ranging from −1 to −4, percentiles 5 and 6 are assigned a score of zero (0), while percentiles 7 to 10 are given positive scores from +1 to +4. The INFLA score comprises four indicators, ranging from −16 to +16. Individuals with higher scores exhibit stronger chronic low-grade inflammation^26^.

### Assessment of genetic susceptibility

The UK Biobank released polygenic risk scores (PRS) of a number of illnesses in September 2022, including the standard PRS for IS^27^. The individuals’ genetic susceptibility was evaluated using the standard PRS for IS (https://biobank.ndph.ox.ac.uk/showcase/field.cgi?id=26248) in this investigation. The IS-PRS utilized in this study was derived by integrating findings from various external genome-wide association studies (GWAS). The PRS can be better understood from prior studies^27,28^. As with previous research^29^, low (the lowest quintile), medium (quintiles 2-4), and high genetic susceptibility (the highest quintile) were the participant classifications used in this study

### Covariates

This study adjusted for sociodemographic potential confounders based on prior research^13,20^, comprising sex (male or female), race (White or others) and age (continuous). Three prevalent methods for assessing socioeconomic status were the Townsend Deprivation Index (TDI) as a continuous metric, educational attainment (college/university degree or alternatives), and average household income is classified into the following ranges (less than 18,000, 18,000 to 30,999, 31,000 to 51,999, 52,000 to 100,000, and exceeding 100,000). The TDI was calculated using the participant’s residential postal code. A higher score reflects an increased level of socioeconomic deprivation in the participant’s residential area. Continuous laboratory tests were conducted to measure CRP. Medication intake comprised antihypertensive medications (present or absent) and lipid-lowering medications (present or absent). Alcohol consumption (categorized as never, previous, or current) and family history of stroke (indicated as yes or no) were evaluated using questionnaires and interviews. We assessed the genetic susceptibility of participants to IS by utilizing the IS-PRS, categorized as low, moderate, or high, while also considering the top 10 genetic principal components.

### Statistical Analyses

For continuous variables, we used mean (standard deviation) or median (interquartile range), and for categorical variables, we used count (percentage) to describe the baseline characteristics of the participants. Analyses of variance (ANOVA) and chi-square tests were used to compare characteristics.

Cumulative incidence curves of IS for low, medium, and high CVH levels were built using the Kaplan-Meier approach. The differences between these levels were assessed using the log-rank test. Using the Cox proportional hazards model, the relationships between LE8 scores, its behavior, biological subscale scores, and individual LE8 components with IS were evaluated. The hazard ratios (HR) and their 95% CI were then calculated. The Schoenfeld residual test was used to evaluate the proportional hazards assumption, and the findings confirmed that the assumption was met. The research utilized three models: the crude model, lacking any covariate adjustments; Age, sex, and race were taken into account in model 1, while PRS, education level, household income, TDI, use of lipid-lowering and antihypertensive medications, CRP, alcohol use, family history of stroke, and PRS were also taken into account in model 2. Furthermore, we utilized Model 2 to develop restricted cubic splines within a Cox regression framework to search for potential dose-response relationships between the LE8 scores, its subscale scores as continuous variables, and the risk of IS. We identified an optimal model with four knots for enhanced precision and reliability using the Akaike Information Criterion (AIC).

The effect of various inflammatory indicators on the association between LE8 scores and IS was investigated using the “mediation” software in R software. The mediation study modified Model 2, excluding CRP. Finally, we conducted 1000 non-parametric bootstrapping simulations to determine the direct effects, indirect effects, and mediation ratios of inflammatory factors.

The association between PRS levels and the risk of IS was initially evaluated in the joint analysis of CVH and IS genetic susceptibility in order to validate the effectiveness of PRS. Based on PRS and CVH levels, we split the participants into nine groups. The reference group was the high PRS and low CVH group, which allow us establish the HR (95% CI) for the other groups. Furthermore, to examine the interaction between genetic susceptibility for IS and CVH, we categorized our study according to genetic susceptibility groups. Subsequently, the likelihood ratio test was used to evaluate the differences when the interaction term (PRS * CVH) was included to the fully adjusted Cox model. An additive interaction model was employed to evaluate the combined effect of genetic susceptibility and CVH on IS risk. The interaction was quantified by relative excess risk (RERI) and attributable proportion (AP). The 95% CI for the AP and RERI will not include zero if there is an additive interaction^30^.

The robustness of the main analysis was assessed by several sensitivity studies. First, to find out if characteristics like age, gender, race, TDI, alcohol use, education level, and household income affected the relationship between CVH and the risk of IS, subgroup analyses were first conducted. Second, we conducted analyses after applying multiple imputations for participants with missing covariates. Third, to take non-IS deaths into account as competing events, we used Fine and Gray’s competing risk model. Fourth, reverse causation was minimized by excluding participants who experienced IS within the first two years of follow-up. Finally, to reduce the likelihood of recall bias, participants who only self-reported IS were excluded from the study. Every statistical analysis was conducted using R software (version 4.3.3). Statistical significance was established as P < 0.05 (two-sided).

## RESULTS

### Descriptive analysis of baseline characteristics

This study included 202,215 participants with complete research data. Table 1 displays the baseline characteristics of subjects categorized by CVH. With a median follow-up duration of 13.73 years, 2,510 patients experienced IS, yielding an incidence rate of 90.60 per 100,000 person-years. The median age of all participants was 56 years (IQR: 49–62), with 52.25% of them being female. The study population included 11,833 participants (5.85%) with low CVH, 157,264 participants (77.77%) with moderate CVH, and 33,118 participants (16.38%) with high CVH. Participants with high CVH were commonly younger, more likely to be female, had higher average household incomes and level of education, and had lower levels of TDI and CRP than those with low CVH. Table S4 displays the baseline characteristics categorized by IS status.

**Table 1.**
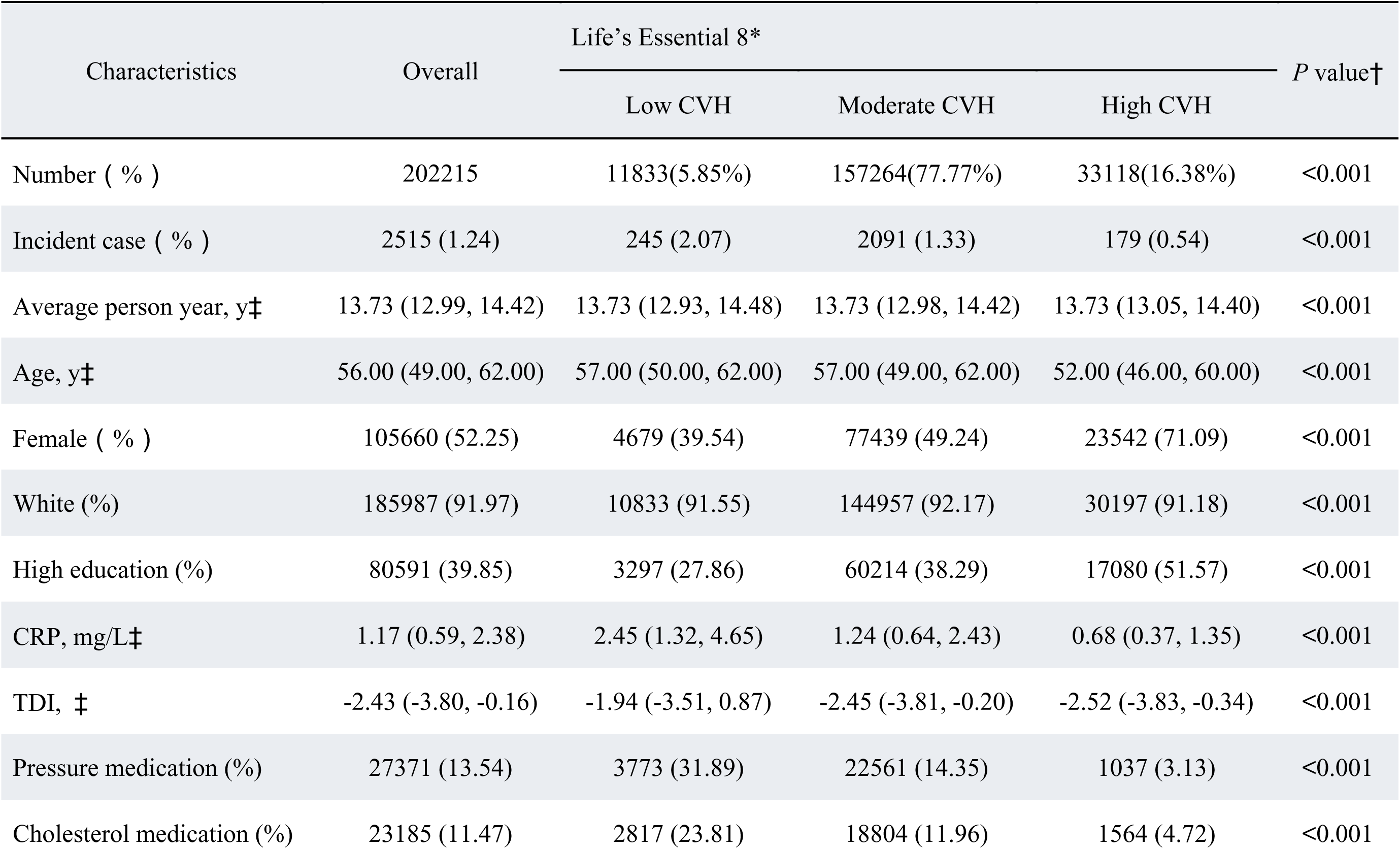
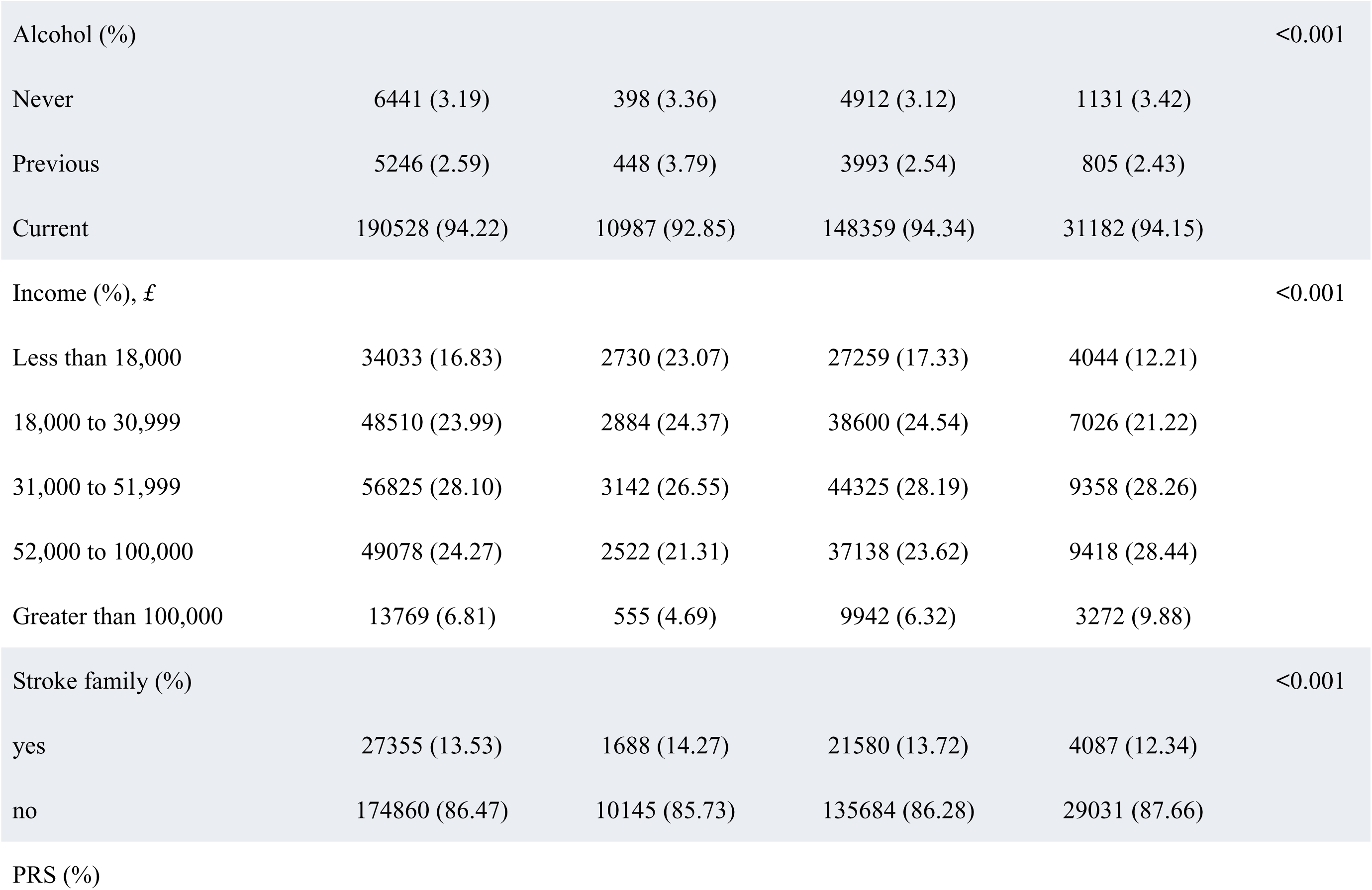
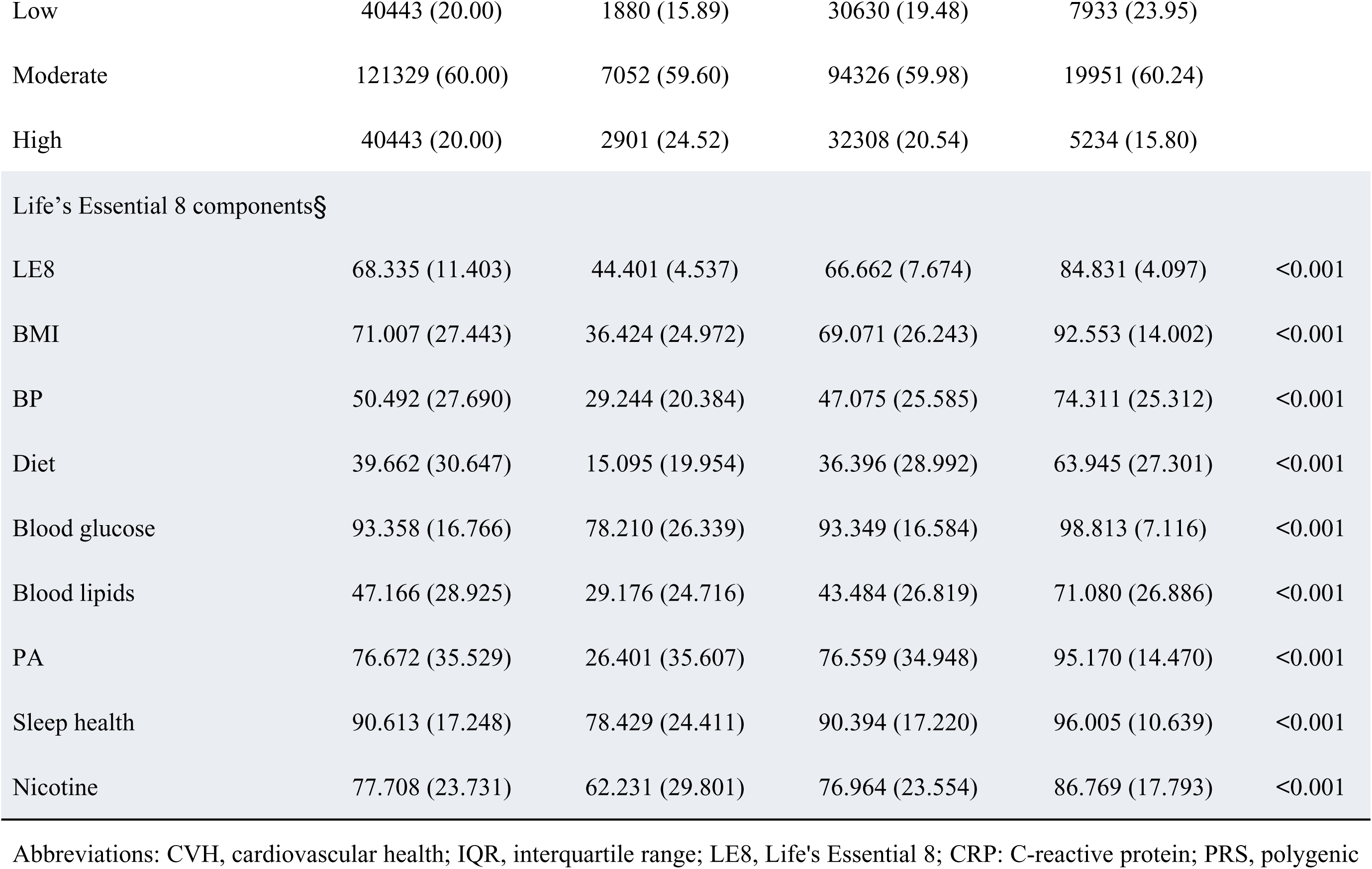
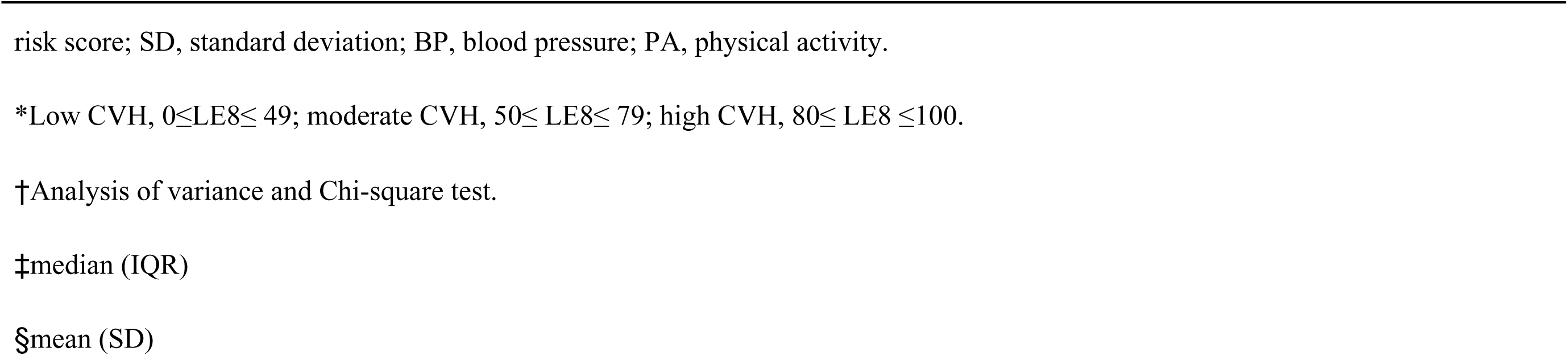
Baseline characteristic of included participants according to LE8 score.

### Association between LE8 and IS risk

Throughout the follow-up period, the cumulative incidence of IS was evaluated based on CVH categories (log-rank tests < 0.0001). Of the three categories, those with low CVH had the greatest cumulative incidence of IS, and moderate and high CVH groups followed. (Figure S2A). The behavior and biological subscale scores presented similar relationships (Figures S2B, C).

The relationship between IS risk and CVH is displayed in Table 2. Individuals with moderate CVH (HR: 0.74; 95% CI: 0.65-0.85) had a 26% lower risk of IS in model 2 (fully adjusted model), whereas those with high CVH (HR: 0.47; 95% CI: 0.39-0.57) had a 53% lower risk compared to those with low CVH. In addition, the rise in continuous variables, including the LE8 scores (HR: 0.84; 95% CI: 0.80-0.87), behavior subscale score (HR: 0.95; 95% CI: 0.91-0.99), and biological subscale score (HR: 0.79; 95% CI: 0.76-0.83), will also decrease the risk of IS. Figure S3 demonstrates that increases in blood pressure, nicotine exposure, sleep, BMI, lipid levels, and blood glucose scores correlate with a decreased IS risk.

**Table 2.**
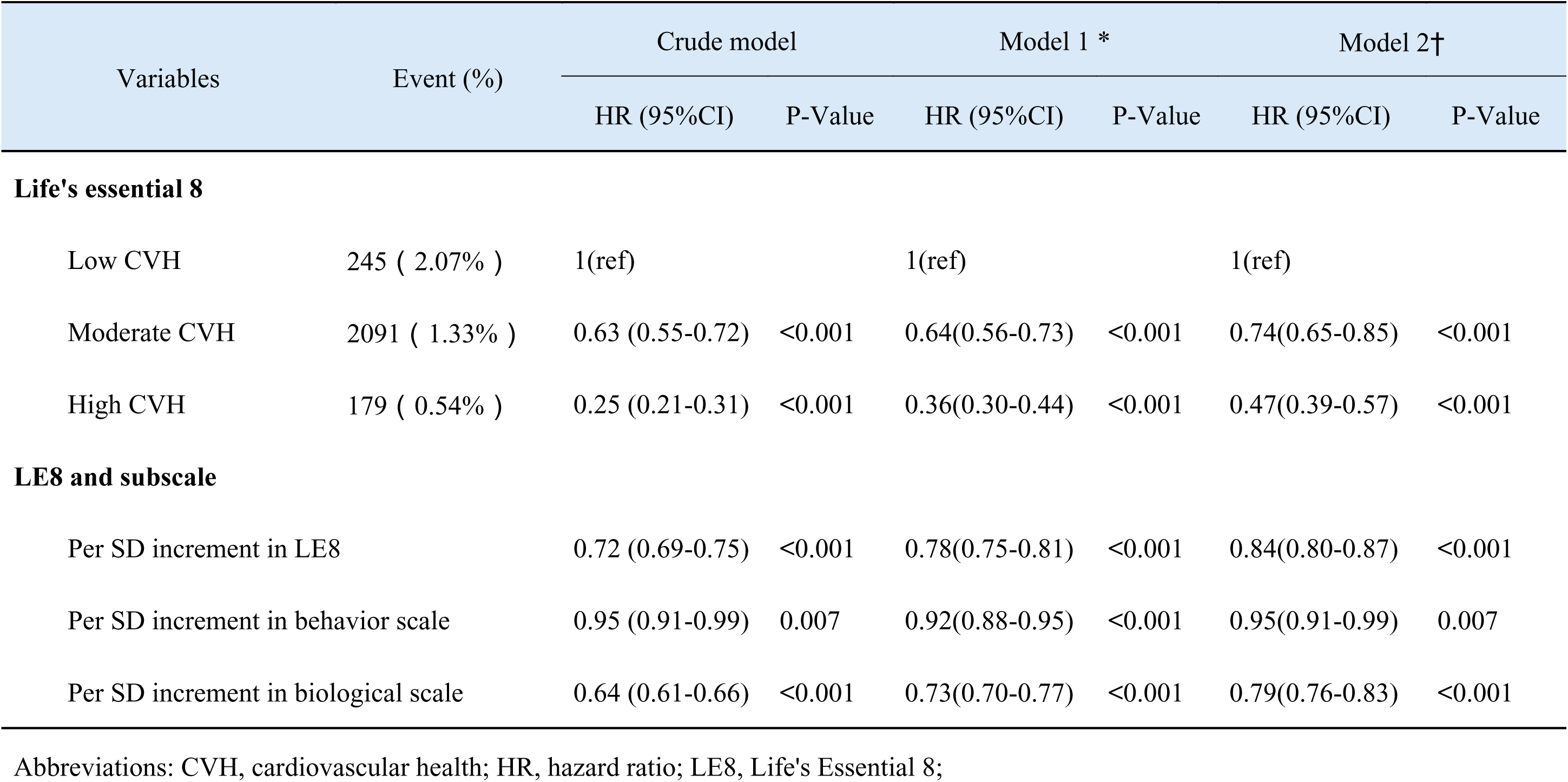
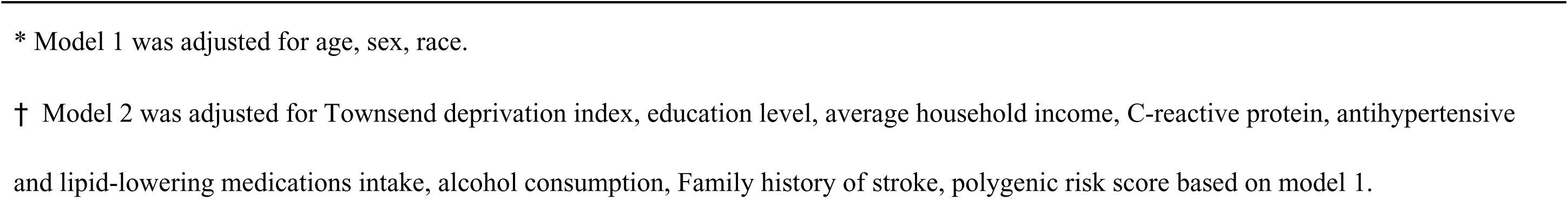
Association of CVH and its subscale scores with the incidence of IS.

Figure 1A illustrates the outcomes of the restricted cubic spline analysis. This analysis revealed a significant negative correlation (*P*-overall < 0.001) and suggested an approximately linear association (*P*-nonlinear = 0.127) between LE8 scores and IS risk. The behavior and biological subscale scores exhibited similar relationships (Figure 1B, C).

**Fig. 1.**
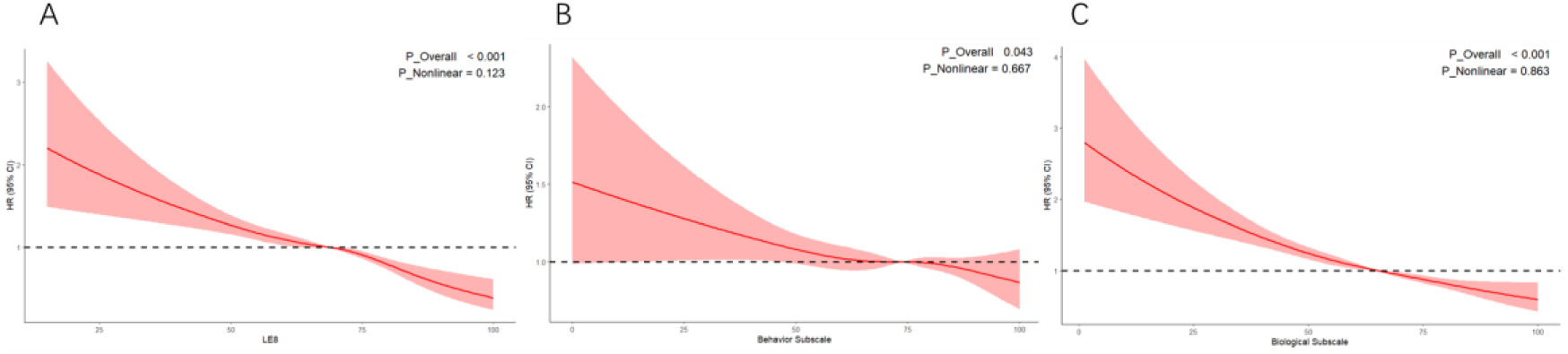
The association of the continuous changes in the (A) Life’s Essential 8score, (B) behavior subscale score, and (C) biological subscale score with the incidence of IS was examined using multivariable Cox regression models (model 2) with restricted cubic splines. CI, confidence interval; HR, hazard ratio; IS, Ischemic Stroke.

### Mediation analysis between LE8 and IS

The mediating effect of inflammatory indicators in the relationship between IS and LE8 scores is depicted in Figure 2. The individual inflammatory markers, including CRP, WBC count, monocytes, platelet count, and eosinophils, played a partial mediating effects between the LE8 scores and IS, with mediation proportions of 12.5%, 4.54%, 3.69%, 3.54%, and 1.91%, respectively. In addition, the composite inflammation index, the INFLA score, also shows similar results to the individual inflammatory markers.

**Figure 2.**
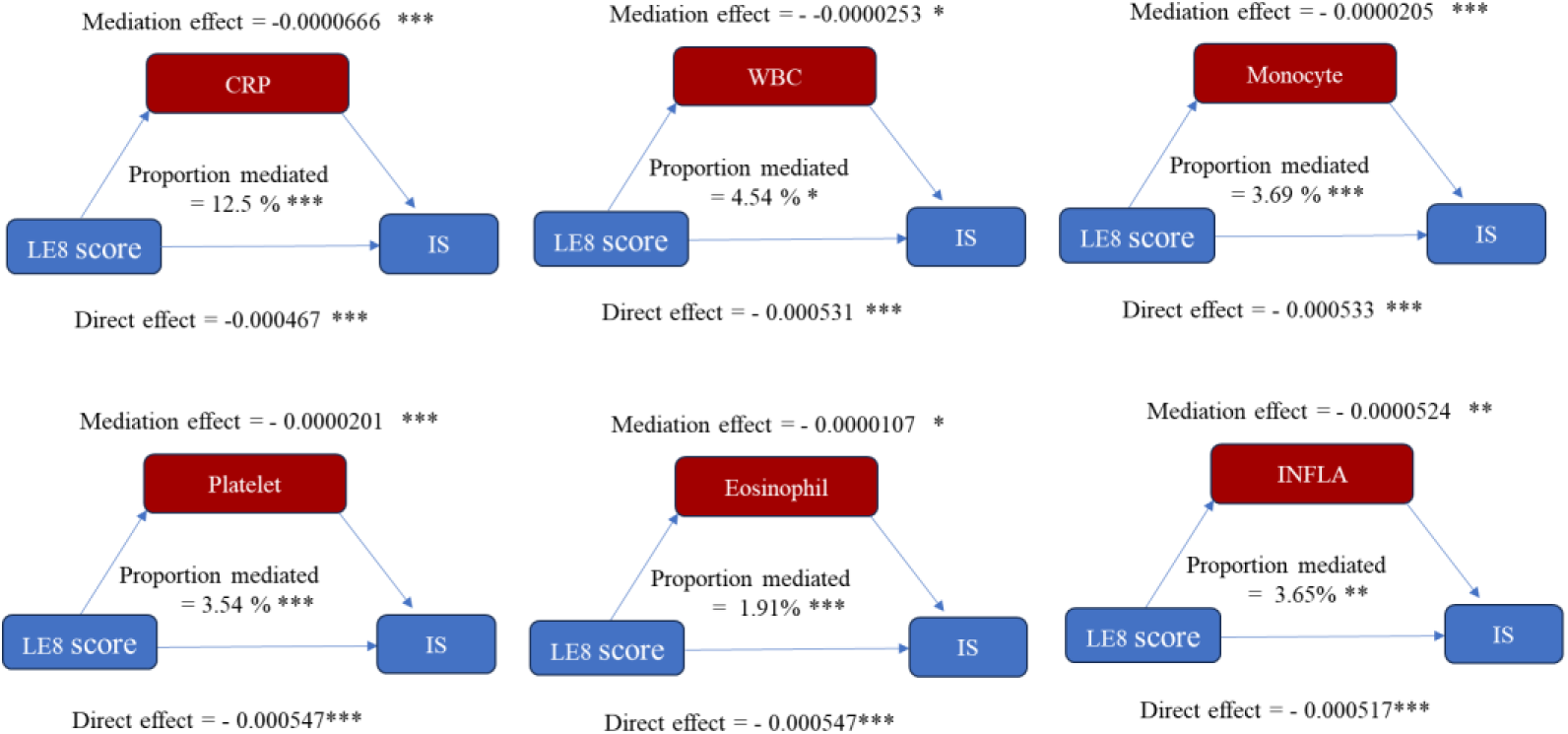
Mediation analyses with inflammatory indicators between the association of CVH and IS (n = 197098). Abbreviations: WBC, white blood cell; CRP, C-reaction protein; IS, Ischemic Stroke; CVH, Cardiovascular Health. Adjusted for age, sex, race, Townsend deprivation index, education level, average household income, antihypertensive and lipid-lowering medications intake, alcohol consumption, Family history of stroke, polygenic risk score. The proportion mediated = Indirect effect / [Indirect effect + Direct effect]. \*\*\**p* < .001; \*\**p* < .01; \**p* < .05.

### Joint and interaction analysis of LE8 and genetic susceptibility

As shown in Table S5, Participants exhibiting higher IS-PRS were more likely to have IS. following adjustment, participants in the moderate genetic susceptibility group (HR: 1.37; 95% CI: 1.28-1.61) exhibited a 37% higher IS risk, while those in the high genetic susceptibility group (HR: 1.98; 95% CI: 1.74-2.25) demonstrated a 98% higher risk compared to the low genetic susceptibility group. Figure 3 illustrates the joint effects between CVH and genetic susceptibility on IS risk. In this joint study, compared to the reference group with high genetic susceptibility and low CVH, the HR was 0.57 (95% CI, 0.43-0.75) for high genetic susceptibility and high CVH and dropped to 0.20 (95% CI, 0.13-0.30) for low genetic susceptibility and high CVH.

**Fig.3.**
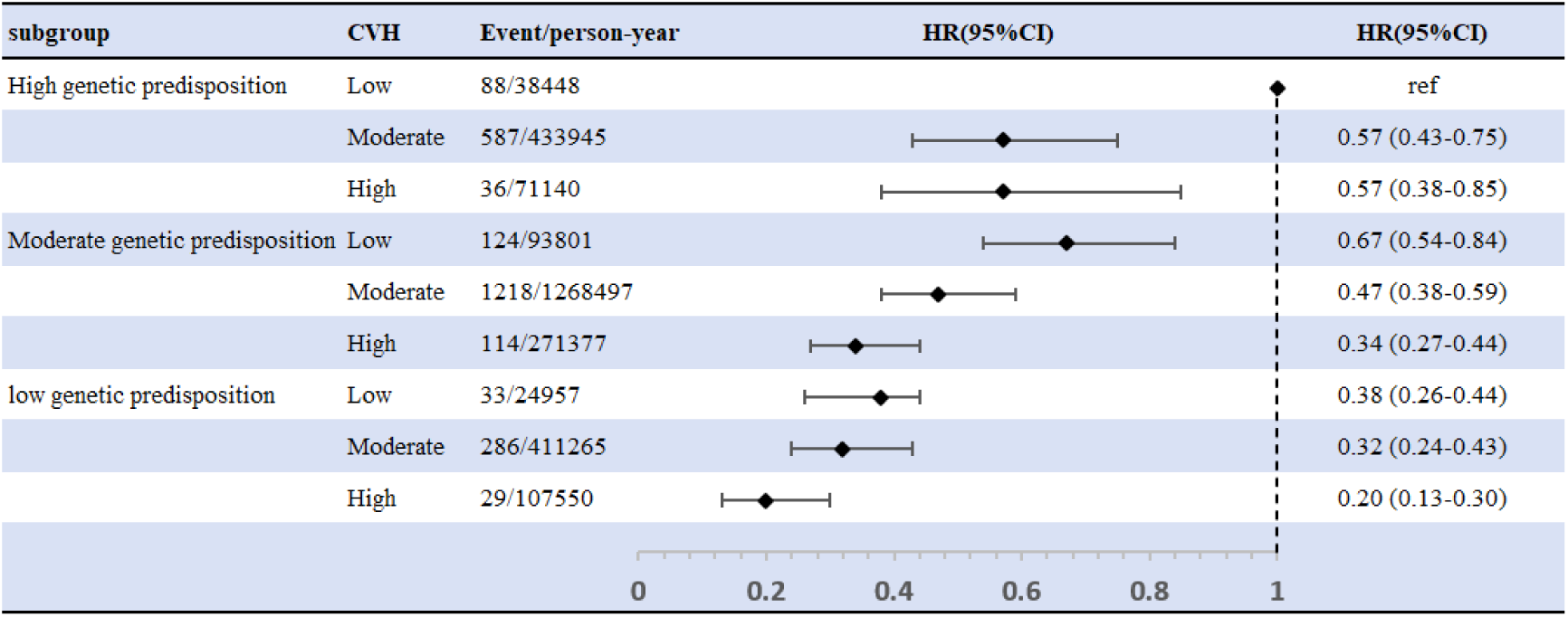
The joint association of PRS and LE8 score on the risk of incident IS. Cox proportional-hazard regression adjusted for age, sex, age, sex, race, drinking status, Townsend deprivation index, education level, average household income, C-reactive protein, antihypertensive and lipid-lowering medications intake, alcohol consumption, Family history of stroke and the top 10 genetic principal components.

In the stratified analysis of genetic susceptibility, all genetic susceptibility levels showed the same negative correlation between IS risk and CVH levels. In contrast to low CVH, for each level of genetic vulnerability, the HRs (95% CIs) for high CVH were 0.35 (0.24-0.53), 0.57 (0.44-0.74), and 0.38 (0.22-0.63). No significant multiplicative interaction (*P* _interaction_ = 0.29) between LE8 and PRS on IS was found (Table 3). Moreover, no significant additive interaction was observed (Table S6).

**Table3.**
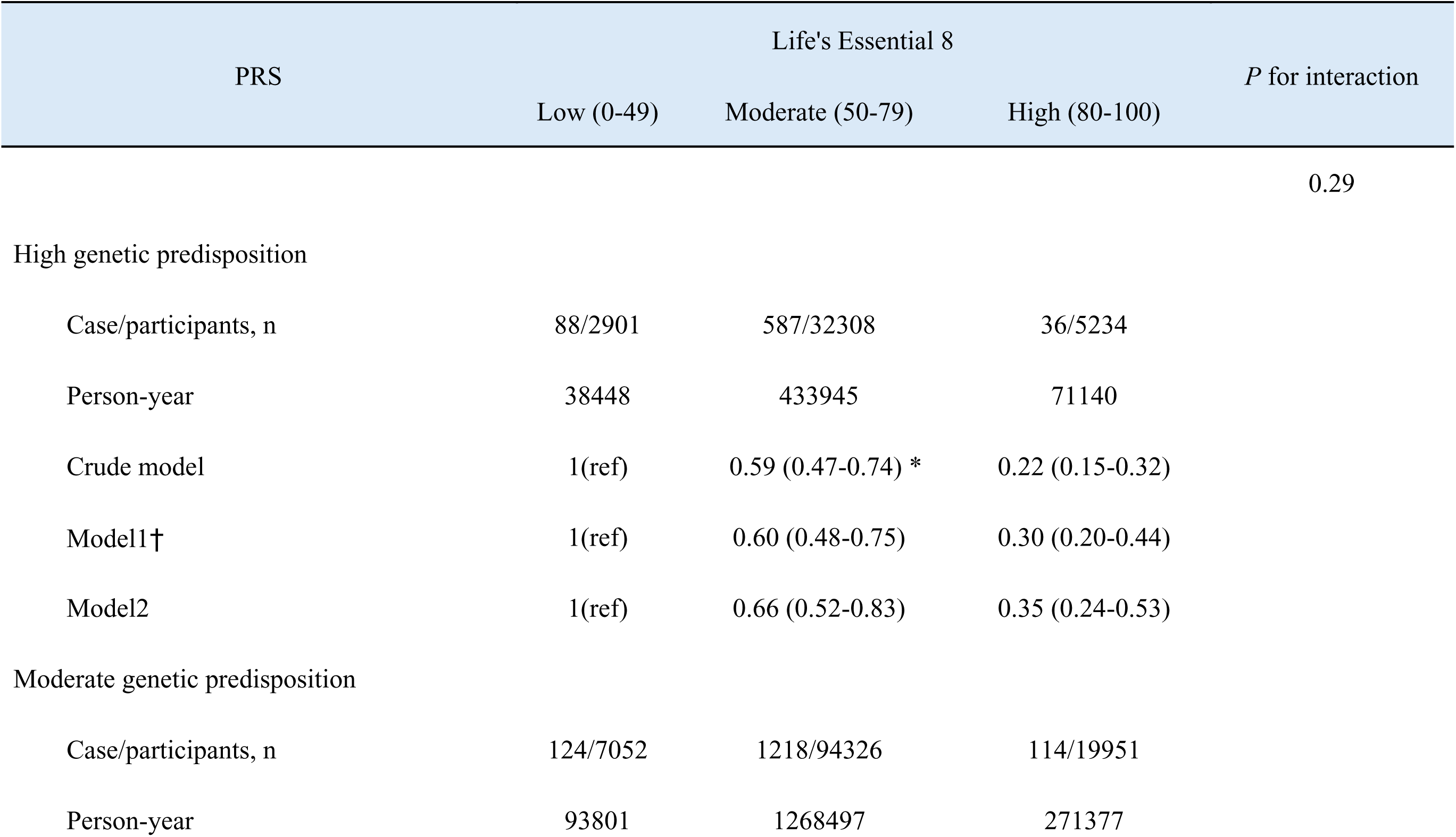
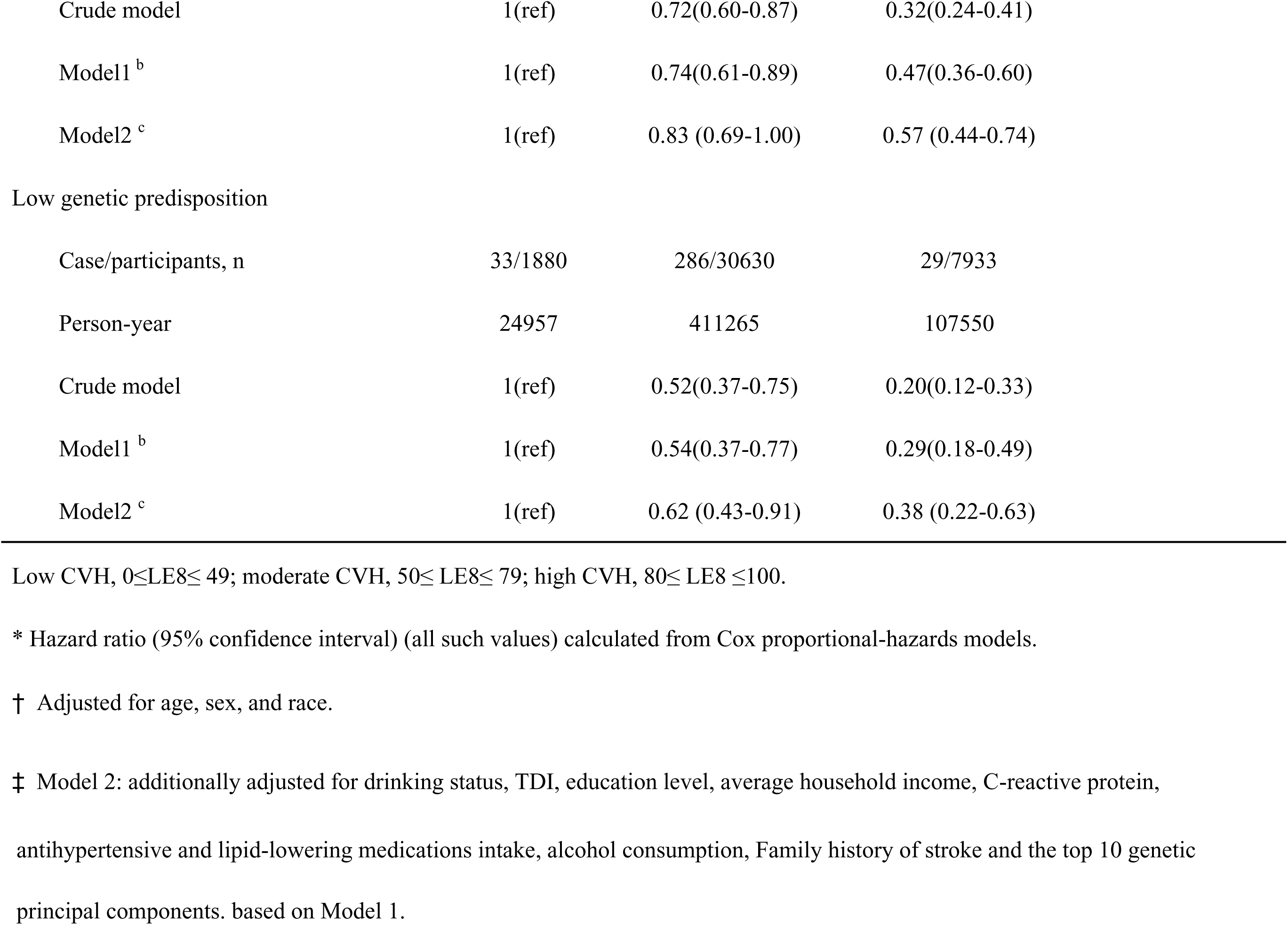
Stratified analysis of association between LE8 score and risk of incident IS according to PRS categories.

| PRS | Life's Essential 8 |  |  | *P* for interaction |
| --- | --- | --- | --- | --- |
|  | Low (0-49) | Moderate (50-79) | High (80-100) |  |
|  |  |  |  | 0.29 |
| High genetic predisposition |  |  |  |  |
| Case/participants, n | 88/2901 | 587/32308 | 36/5234 |  |
| Person-year | 38448 | 433945 | 71140 |  |
| Crude model | 1(ref) | 0.59 (0.47-0.74) * | 0.22 (0.15-0.32) |  |
| Model1† | 1(ref) | 0.60 (0.48-0.75) | 0.30 (0.20-0.44) |  |
| Model2 | 1(ref) | 0.66 (0.52-0.83) | 0.35 (0.24-0.53) |  |
| Moderate genetic predisposition |  |  |  |  |
| Case/participants, n | 124/7052 | 1218/94326 | 114/19951 |  |
| Person-year | 93801 | 1268497 | 271377 |  |

|  |  |  |  |
| --- | --- | --- | --- |
| Crude model | 1(ref) | 0.72(0.60-0.87) | 0.32(0.24-0.41) |
| Model1<sup>b</sup> | 1(ref) | 0.74(0.61-0.89) | 0.47(0.36-0.60) |
| Model2<sup>c</sup> | 1(ref) | 0.83 (0.69-1.00) | 0.57 (0.44-0.74) |
| Low genetic predisposition |  |  |  |
| Case/participants, n | 33/1880 | 286/30630 | 29/7933 |
| Person-year | 24957 | 411265 | 107550 |
| Crude model | 1(ref) | 0.52(0.37-0.75) | 0.20(0.12-0.33) |
| Model1<sup>b</sup> | 1(ref) | 0.54(0.37-0.77) | 0.29(0.18-0.49) |
| Model2<sup>c</sup> | 1(ref) | 0.62 (0.43-0.91) | 0.38 (0.22-0.63) |

Low CVH, $0 \leq \text{LE8} \leq 49$ ; moderate CVH, $50 \leq \text{LE8} \leq 79$ ; high CVH, $80 \leq \text{LE8} \leq 100$ .
\* Hazard ratio (95% confidence interval) (all such values) calculated from Cox proportional-hazards models.
† Adjusted for age, sex, and race.
‡ Model 2: additionally adjusted for drinking status, TDI, education level, average household income, C-reactive protein,
antihypertensive and lipid-lowering medications intake, alcohol consumption, Family history of stroke and the top 10 genetic principal components. based on Model 1.

### Sensitivity analyses

Our findings were subjected to numerous sensitivity analyses to guarantee their robustness. First, we conducted further subgroup analyses, stratified by potential risk variables including sex, age, race, and average household income and education level. All of these subgroup analyses showed the same link among CVH and IS (Table S7). Interestingly, Participants with high CVH levels before 60 years of age had a considerably decreased risk of IS than those over 60 at baseline (*P* _for interaction_ < 0.001). Similarly, compared to white participants, participants of other races exhibited lower IS risk (*P* _for interaction_ = 0.041). Other subgroups did not exhibit any significant interactions (*P* for interaction > 0.05).

Secondly, We conducted the following sensitivity analyses: (1) Individuals who experienced IS during the initial two years of monitoring were excluded (Table S8); (2) Using Fine and Gray competing risk analysis to evaluate the competitive risk of non-IS death (Table S9); (3) Employing multiple imputation to account for participants previously excluded due to missing covariates (Table S10); (4) excluding individuals who only self-reported having IS (Table S11).

## DISCUSSION

In this study, we found that increased LE8 scores are significantly associated with a reduced risk of IS, according to this large-scale prospective cohort research, with inflammation partially mediating this association. Participants with high CVH exhibited a 53% lower incidence of IS than those with low CVH. This association remains stable across various subgroup analyses and sensitivity analyses. Furthermore, the protective relationship of CVH and IS risk may be strengthened in cases of lower genetic susceptibility.

The most recent CVH score suggestion made by the AHA is LE8. In comparison to the prior LS7 system, LE8 has undergone several improvements^9^. The correlation between CVH and IS in the LS7 assessment is well established^10–12^; however, research on the relationship between CVH and IS in the updated LE8 assessment remains limited and presents inconsistent results. A cohort study conducted in Kailuan, China, with 68,854 participants, found a significant inverse relationship between IS risk and the CVH as assessed by the LE8 scores^13^. This aligns with the conclusions that we obtained from our investigation. Another study involving 2,269 American Indians found no statistical correlation between LE8-assessed CVH and the incidence of new IS^14^. The observed difference may be attributed to the study’s failure to document sleep duration, resulting in the overall score being derived from only 7 of the 8 AHA LE8 indicators. This may cause the LE8 scores to not match the actual situation. The restricted number of IS events, small number of participants, and short follow-up period of the study might help to explain its inability to detect a correlation. Our ongoing UK biobank cohort study not only found a significant inverse relationship among LE8 scores, its subscale scores, CVH levels, and IS risk. Additionally, we observed the dose-response correlation between them and found that the scores of the two subscales, the LE8 scores, and IS risk have an approximately linear relationship. Although rapid reperfusion through intravenous thrombolysis and endovascular thrombectomy has been shown to reduce the disability rate of IS^31,32^, this study’s findings provide new insights for future preventive and interventional treatments.

This study is, to our knowledge, the first evidence of the combined effect of genetic susceptibility and CVH, as assessed by LE8, on the risk of IS. According to our findings, compared to the reference group characterized by low CVH and high genetic susceptibility, the group exhibiting high CVH and low genetic propensity has an 80% decreased risk of incident IS. Moreover, in terms of the risk of IS, there was no evidence of any interaction between CVH and genetic susceptibility. Previous research has mostly concentrated on how genetic predisposition and a healthy lifestyle interact with IS^20,21^. According to two research, a healthy lifestyle and genetic predisposition may interact. Adopting a healthy lifestyle could reduce or partially offset IS risk associated with genetic susceptibility. Two studies on LE8 and stroke show that there is no interaction between LE8 and genetic susceptibility to stroke risk^33,34^. This is in line with the conclusions that we obtained from our research on IS. It’s interesting to observe that this study’s stratified and interaction analyses revealed that CVH significantly reduces IS, and that this protective impact is stronger in those who have lower genetic susceptibility. This suggests that populations with lower genetic susceptibility also need to sustain high levels of CVH to improve the prevention of IS. In the context of public health, clarifying the interplay between CVH and IS genetic factors may guide primary prevention strategies for IS.

To comprehend the relationship between LE8 and IS deeper, it is crucial to explore its potential mechanisms. The components in LE8 may decrease the occurrence of IS through several mechanisms. First, chronic inflammation may significantly contribute to this process. Evidence indicates that smokers exhibit a more robust inflammatory response compared to non-smokers, a phenomenon that is especially evident in patients with IS^35^. By reducing inflammatory responses and improving vascular function, a balanced diet—especially one rich in fruits and vegetables that are high in antioxidants—can lower the risk of cardiovascular illnesses^36^. The inflammatory state accelerates endothelial cell damage and dysfunction while increasing vascular wall permeability, facilitating the entry of harmful substances into the vascular wall^37^. The inflammatory response activates immune cells that are functioning abnormally. These cells accumulate in damaged blood vessels, exacerbating the injury through the release of inflammatory factors^37^. This series of processes contributes to atherosclerosis development and markedly elevates the risk of IS^38^. The results of this research confirmed the mediating role of inflammatory factors and demonstrated that LE8 can effectively safeguard the population from IS damage by lowering the levels of various inflammation-related factors. Second, oxidative stress may be linked to inflammatory responses as a biological mechanism connecting LE8 and IS^39^. Exercise demonstrates an antioxidant effect through the inhibition of inflammatory pathways, resulting in various physiological advantages for CVH^40^. Excessive production of reactive oxygen species (ROS) can occur when blood pressure, glucose, and cholesterol levels are not maintained within healthy ranges^41^. This leads to heightened oxidative stress and inflammation, which aggravates vascular damage and increases risk. Consequently, it is crucial to further investigate the mechanisms behind the relationship between LE8 and IS.

The study’s primary advantages are its prospective design, which includes an extensive follow-up period with subjects and a large number of samples gathered from the UK Biobank, which offers sufficient outcome events and statistical capabilities. Mediation analysis and interaction analysis are used to clarify the intrinsic mechanisms of research factors. The stability of the results is guaranteed by several sensitivity assessments. However, this research has several limitations. First, all eight LE8 indicators used in this study were obtained by self-report, potentially resulting in recall bias and misclassification bias. Furthermore, it is difficult to assess how longitudinal changes in the LE8 indicators affect IS risk because all variables were measured at baseline. Second, because white British people made up the majority of participants in the UK Biobank database, the results might not be applicable to other ethnic groups. Third, while various potential confounders were accounted for, the influence of residual confounders could not be entirely eliminated. Fourth, as this study is a volunteer cohort, participants are likely to exhibit greater health awareness and typically possess higher education levels and socioeconomic status, potentially resulting in volunteer bias^42^. Fifth, covariates with missing data were excluded from the study, leading to a reduced sample size. Nonetheless, the results demonstrated that the relationships were constant when we completed in the missing data using multiple imputation.

### Conclusions

In conclusion, the current research shows that LE8 scores, behavior subscale score, biological subscale score, and CVH are all significantly associated with IS risk. Systemic inflammation plays a role in partially mediating the association between LE8 scores and IS risk. Additionally, these protective relationships can be strengthened in cases of lower genetic susceptibility. These findings provide valuable insights into the etiology of IS, which can help advance disease prevention and control efforts.

## Data Availability

Data availability The data supporting the findings of this study are available in the UK Biobank repository. Accessible at: https://www.ukbiobank.ac.uk.

## Non-standard Abbreviations and Acronyms

IS: ischemic stroke
CVH: cardiovascular health
AHA: American Heart Association
LS7: Life’s Simple 7
non-HDL: non-high-density lipoprotein
CRP: C-reactive protein
ICD-10: tenth International Classification of Diseases
WBC: white blood cell
GrL: granulocyte/lymphocyte ratio
PRS: polygenic risk scores
GWAS: genome-wide association study
AIC: Akaike Information Criterion
RERI: relative excess risk
AP: attributable proportion
ROS: reactive oxygen species

## Acknowledgments

The authors thank the UK Biobank personnel and all study participants. Under Application Number 95715, the UKB Resource was used to conduct this study.

## Funding

There was not any external financing for this study.

## Disclosures

None.

## Notes

### Competing Interest Statement

The authors have declared no competing interest.

### Author Declarations

The North-West Multi-Centre Research Ethics Committee approved the UK Biobank study.

